# A case of SARS-CoV-2 carrier for 32 days with several times false negative nucleic acid tests

**DOI:** 10.1101/2020.03.31.20045401

**Authors:** Lingjie Song, Guibao Xiao, Xianqin Zhang, Zhan Gao, Shixia Sun, Lin Zhang, Youjun Feng, Guangxin Luan, Sheng Lin, Miao He, Xu Jia

**Affiliations:** Non-coding RNA and Drug Discovery Key Laboratory of Sichuan Province, Chengdu Medical College, Chengdu, Sichuan, China; The First People’s Hospital of Ziyang City, Ziyang, Sichuan, China; Basic Medical College, Chengdu Medical College, Chengdu, Sichuan, China; Institute of Blood Transfusion, Chinese Academy of Medical Sciences, Chengdu, Sichuan, China; Department of Pharmacy, Shaoxing People’s Hospital; Shaoxing Hospital, Zhejiang University School of Medicine, Shaoxing, Zhejiang, China; Department of Medical Microbiology and Parasitology, Zhejiang University, Hangzhou, China

**Author notes:** Correspondence to: Xu Jia; Miao He; Sheng Lin. These authors contribute equally to this article (Lingjie Song, Guibao Xiao).

**Keywords:** SARS-CoV-2, COVID-19, RT-PCR, falsely negative

## Abstract

In 2019, a novel coronavirus (SARS-CoV-2) was first discovered in Wuhan, Hubei, China, causing severe respiratory disease in humans, and has been identified as a public health emergency of international concern. With the spread of the virus, there are more and more false negative cases of RT-PCR nucleic acid detection in the early stage of potential infection. In this paper, we collected the epidemiological history, clinical manifestations, outcomes, laboratory results and images of a SARS-CoV-2 carrier with no significant past medical history. The patient was quarantined because of her colleague had been diagnosed. After the onset of clinical symptoms, chest CT results showed patchy ground-glass opacity (GGO) in her lungs, but it took a total of nine nucleic acid tests to confirm the diagnosis, among which the first eight RT-PCR results were negative or single-target positive. In addition to coughing up phlegm during her stay in the hospital, she did not develop chills, fever, abdominal pain, diarrhea and other clinical symptoms. Since initial antiviral treatment, the lung lesions were absorbed. But the sputum nucleic acid test was still positive. In combination with antiviral and immune therapy, the patient tested negative for the virus. Notably, SARS-CoV-2 was detected only in the lower respiratory tract samples (sputum) throughout the diagnosis and treatment period. This is a confirmed case of SARS-CoV-2 infection with common symptoms, and her diagnosis has undergone multiple false negatives, suggesting that it is difficult to identify certain carriers of the virus and that such patients may also increase the spread of the SARS-CoV-2.

## Introduction

Since the outbreak of severe acute respiratory syndrome (SARS) coronavirus 2 (SARS-CoV-2) in Wuhan (Hubei, China) in December 2019 [1], it has spread rapidly across the globe and has become a major public health emergency worldwide. The international commission classification of viruses (ICTV) announced that 2019-nCoV was officially classified as SARS-CoV-2[2]. The world health organization (WHO) announced that the official name of the disease caused by this virus is coronavirus disease (COVID-19) [3]. The outbreak has led the Chinese government to take drastic measures to contain the outbreak, including isolating millions of residents in Wuhan and other affected cities. According to the “Diagnosis and Treatment of COVID-19 (Trial Version 7)”, the disease must be confirmed by reverse transcription polymerase chain reaction (RT-PCR) or pathogen gene sequencing from respiratory, digestive and blood samples. The RT-PCR test is often used for clinical detection. Recently, it has been reported that the patients had clinical symptoms but their nasopharyngeal swab showed negative RT-PCR results[4]. Thus, it is necessary to study the causes of these multiple RT-PCR negative results.

In order to better diagnose COVID-19, this study analyzed a case whose eight RT-PCR tests were negative. According to the epidemiological history and clinical data of the case, this study aimed to better diagnose the incubation period and asymptomatic patients by combining different diagnostic methods, thus reducing the spread of the disease.

## Methods

This study investigated a case of highly suspected SARS-CoV-2 infection admitted to a hospital. The results of RT-PCR were negative for eight times and positive for the ninth time.

### Data Collection

The epidemiological and clinical data from the information system of the hospital, including possible exposure to pathogens, visits to health facilities, hospitalization, treatment, pathogens and laboratory tests, and clinical results etc. were collected by an investigator. The records were verified by another investigator. A third researcher further verified the data by cross-checking the information in the medical records system and attending physician.

The original image of chest computed tomography (CT) was provided by the local county people’s hospital.

### SARS-CoV-2 Real-Time Quantitative PCR

According to the guidelines released by the National Health Commission of People’s Republic of China (NHC of China), nucleic acid testing kits were used. The samples were throat swabs, sputum, anal swabs, feces and plasma. The detection kit simultaneously detected three target genes of SARS-CoV-2, including RdRp, envelop protein (E) and nucleocapsid protein (N).

## Results

### Epidemiological information

The reported case was a middle-aged woman who worked in a hotel and returned home on January 23. She has not been to any other places since returning home, has no contact with people returning from the affected areas, has no travel history to Wuhan and has denied any contact with wild animals. The patient had been admitted to a local health clinic for coughing after a cold. After her colleagues were diagnosed with COVID-19, the patient was transferred to the local county people’s Hospital for isolation and observation on February 2. It was later learned that about 8 colleagues were diagnosed with COVID-19.

During intensive medical observation on February 2 to February 6, the patient showed no other clinical symptoms. The tests of nucleic acid from pharyngeal swab were negative on February 7, 8, 11 and 13. But the CT reports of her lung (Fig 1) on February 12 and 15 showed patchy ground-glass opacity (GGO) in her lung, indicating that the patient was highly suspected of SARS-CoV-2 infection. Subsequently, the results of two RT-PCR tests were negative.

**Figure 1:**
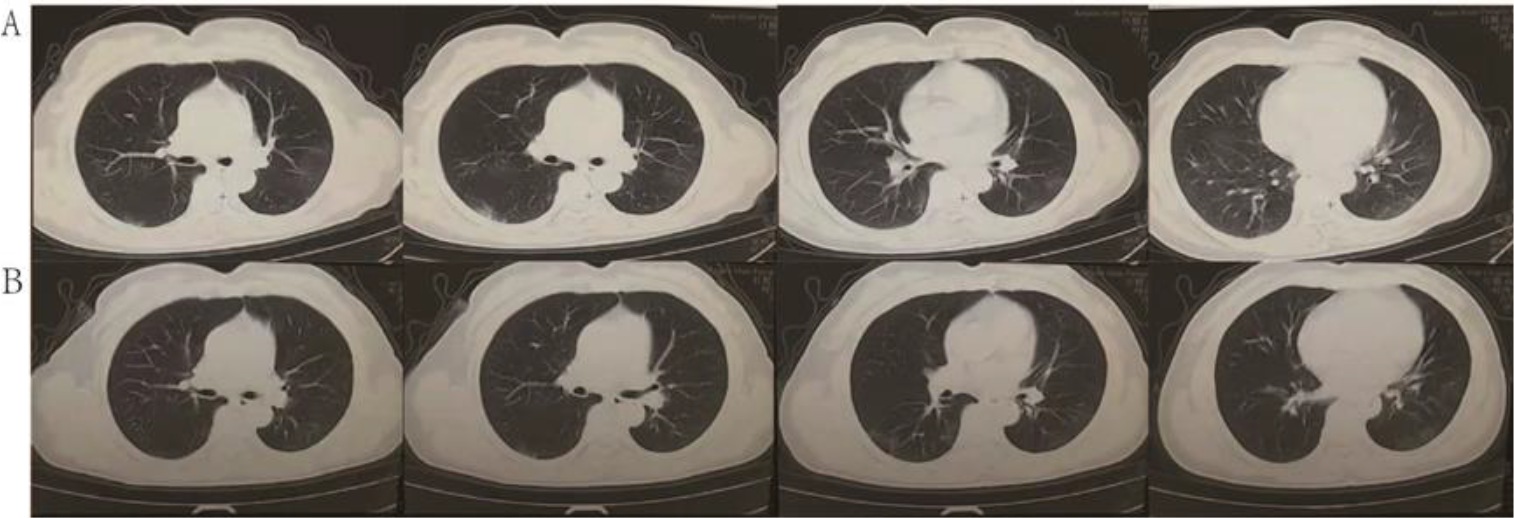
The patient’s Chest CT images. A: CT images on Feb 12; B: CT images on Feb 15.

In order to further improve the relevant examination and treatment, the patient was transferred to our hospital on February 21 for further isolation treatment. The tests of nucleic acid were still negative on February 22 and single-target positive on February 23. Until February 24, the ninth practical sputum nucleic acid test was positive, and the hospital confirmed COVID-19 infection to her clinical symptoms and laboratory test results. Nucleic acid tests were then performed on those who were in close contact with the patient. However, all results were negative.

### Clinical information and follow-ups

This patient had no chronic history of hypertension, diabetes, coronary heart disease, etc. At the time of admission, the patient presented acute symptoms, with body temperature at 36.6°C, pulse rate at 78 beats per minute, respiratory rate at 20 beats per minute, blood pressure of 136/79mmHg, and subcutaneous oxygen saturation (SpO2) of 98.6%. No other abnormalities were detected in pneumonia-associated nucleic acid tests and respiratory pathogenic nucleic acid tests. The remaining laboratory results were shown in table 1. At the time of admission, the patient had been coughing for more than 20 days and had no symptoms of dyspnea, diarrhea or vomiting. There were no clinical symptoms such as fever, diarrhea and vomiting except cough during the hospitalization. The body temperature changes between admission and February 25 were shown in Fig 2.

**Table 1.** Laboratory test of the patient.

|  | Feb 12 | Feb 20 | Feb 22 | Reference range |
| --- | --- | --- | --- | --- |
| WBC, $\times 10^9/L$ | 6.97 | 5.23 | 4.74 | 3.5-9.5 |
| RBC, $\times 10^{12}/L$ | 3.77 | 3.6 | 3.63 | 3.8-5.8 |
| Neutrophil, % | 84.3 | 48.1 | 63.5 | 40-75 |
| Lymphocyte, % | 11.5 | 39.4 | 27.4 | 20-50 |
| Hemoglobin, g/L | 125 | 123 | 120 | 115-150 |
| PLT, $\times 10^9/L$ | 240 | 205 | 205 | 125-350 |
| PT, s | N | N | 12.3 | N/8.8-13.8 |
| APTT, s | N | N | 31.7 | N/28-42 |
| ALT, IU/L | N | N | 10 | N/7-40 |
| AST, IU/L | N | N | 17 | N/13-35 |
| Total bilirubin, mmol/L | N | N | 18 | N/2-20.4 |
| Potassium, mmol/L | N | N | 3.81 | N/3.5-5.3 |
| Sodium, mmol/L | N | N | 142.7 | N/137-147 |
| Creatinine, umol/L | N | N | 47 | N/44-106 |
| BUN, mmol/L | N | N | 2.36 | N/2.6-7.5 |
| CRP, mg/L |  |  | 0.3 | 0-10 |
| Creatine kinase, U/L |  |  | 43 | 40-200 |
| CKMB, U/L | N | N | 7 | N/<24 |

Abbreviations: WBC, white blood cell count; RBC, red blood cell; PT, prothrombin time; APTT, activated partial thromboplastin time; ALT, alanine aminotransferase; AST, aspartate aminotransferase; BUN, blood urea nitrate; CKMB, creatine kinase-MB; N, normal.

**Figure 2:**
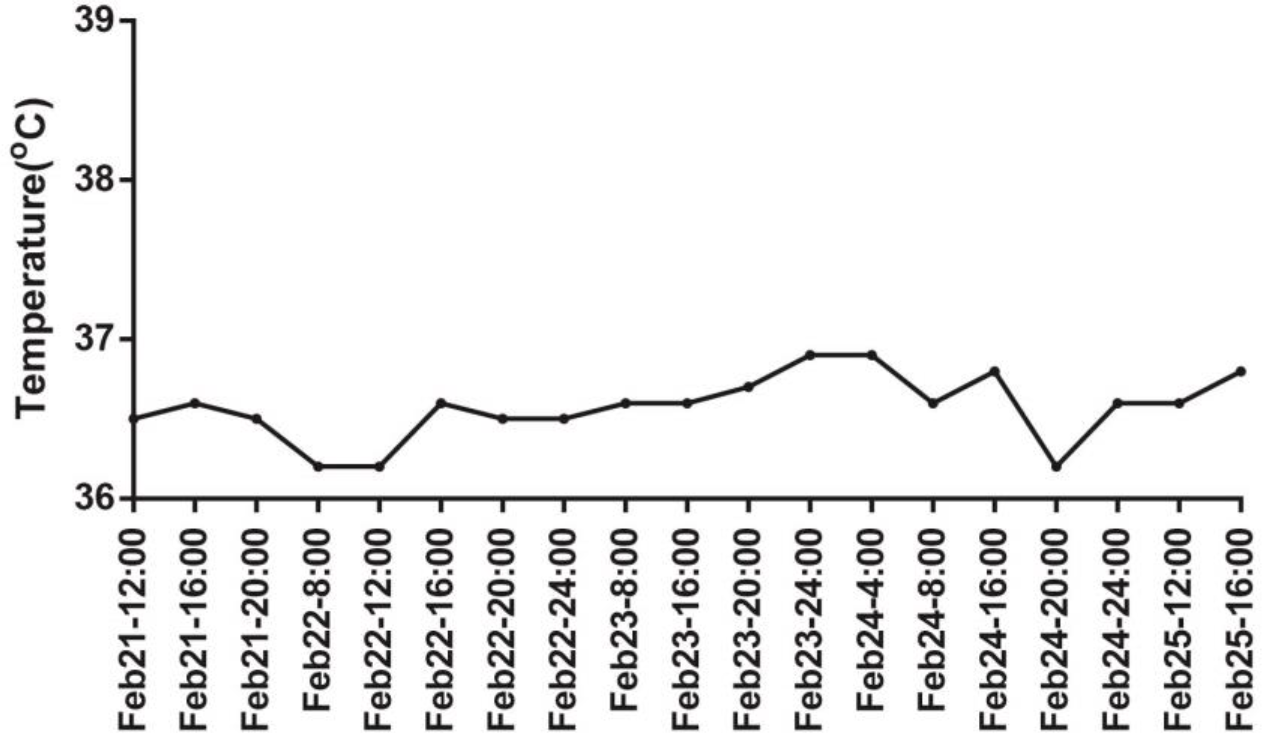
Body temperature of the patient during hospitalization between February 21 – 24, 2020.

Arbidol tablets were given on the day of admission for 10 consecutive days and chloroquine phosphate tablets were continuous used for 8 days. Imaging examination showed that the lung lesions were absorbed before comparison (Fig3). But the nucleic acid test of sputum fluid was still positive. On March 3, arbidol tablets were discontinued and lopinavir/ritonavir tablets were used instead. Meantime she was treated with aerosolized interferon α1β to continue antiviral treatment. Combined with moxifloxacin antibacterial therapy and immunotherapy, the patient’s symptoms were alleviated.

**Figure 3:**
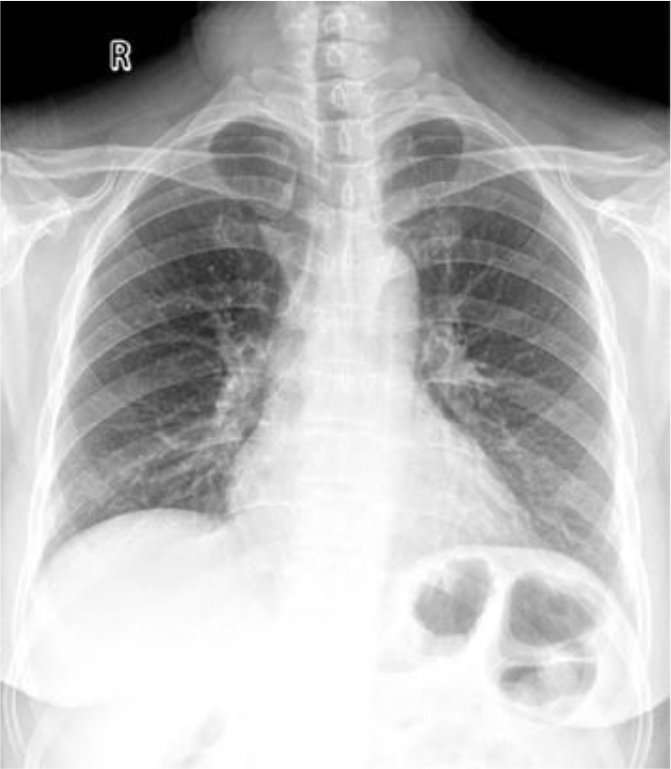
Chest Radiographs image of the patien on Feb 28. After treatment, lesion in the lungs were reduced. Hardly GGO samples were seen.

After 20 days hospitalization, the patient’s physical examination showed good results that the vital signs were normal, clear in mind, mentally acceptable, no dry and wet rales in both lungs, no positive signs were found in heart and abdomen, and the pathological signs were negative. Although the patients still had occasional cough and sputum, a small amount of white foam-like sputum, fearless of cold, fever, dyspnea, but had no abdominal pain and diarrhea, indicating that the general condition of the patient was normal. After discussion by the expert group, the patient met the discharge standards. She was discharged on March 12.

### SARS-CoV-2 nucleic acid test results

Since admission on February 21, a total of eleven SARS-CoV-2 nucleic acid tests have been performed on the patient. The samples that tested positive were sputum, and the rest were negative. The specific test results were shown in Table 2.

**Table 2.** RT-PCR results of different samples.

|  | Feb 22 | Feb 23 | Feb 24 | Feb 29 | Mar 1 | Mar 2 | Mar 4 | Mar 7 | Mar 8 | Mar 10 | Mar 11 |
| --- | --- | --- | --- | --- | --- | --- | --- | --- | --- | --- | --- |
| OP swab | - |  |  |  |  |  |  |  | - | - | - |
| sputum |  | N+/E+ | + | - | + | + | + | - | + | - | - |
| anal swab | - |  |  | - |  |  |  | - | - |  | - |
| excrement | - |  |  |  |  |  |  |  | - |  | - |
| plasma | - |  |  | - |  |  |  |  |  |  |  |

Abbreviations: OP, oropharyngeal; —, negative; +, positive; N, nucleocapsid protein; E, envelop protein.

## Discussion

This article reported a case of SARS-CoV-2 infection that was negative for eight RT-PCR tests. The patient has had close contact with a confirmed patient and belongs to the third or fourth generation of transmission patients [5]. Other than cough, there were no other clinical symptoms such as fever, fatigue, muscle pain, and the infection status was hidden at early stage. The patient has been infected for at least 32 days, and a total of eight RT-PCR tests were negative or single-target positive before the diagnosis. The number of tests was more than the current report and took a long time. Although the nucleic acid test was negative or single-target positive, the low number of white blood cells and lymphocytes in laboratory tests, and GGO in the lungs by CT examination indicated SARS-CoV-2 infection. It can be seen that using RT-PCR as a nucleic acid test cannot completely rule out SARS-CoV-2 infection. There are several factors that may cause the negative results when troubleshooting sampling and transportation issues.

First, sample collection. Same as SARS-CoV[6,7], the functional receptor targeted by SARS-CoV-2 is angiotensin-converting enzyme 2 (ACE2) [8]. ACE2 is found mainly in type I epithelial cells and type II epithelial cells in the alveoli, and it is less abundant in cells such as airway epithelial cells, fibroblasts, endothelial cells and macrophages, so the amount of virus in the lower respiratory tract is significantly higher than that in the upper respiratory tract. Due to the reason that RT-PCR detection requires the cells containing a sufficient amount of virus, the virus-containing cells must be sampled when collecting specimens. Recent studies have shown that the viral load in sputum was greater than that in throat swabs[9]. The patient’s RT-PCR positive nucleic acid samples during the hospitalization were all deep sputum collected after being atomized by 3% sodium chloride injection. Combined with the reported cases [10], it is suggested that the cause of several negative nucleic acid tests in highly suspected patients may be related to the sample collection site.

Compared with SARS-CoV, SARS-CoV-2 has a higher load in the upper respiratory tract [11], and the virus in the upper respiratory tract is more likely to fall off and spread, which may make it easier spread when asymptomatic. However, in the case of the patient, no virus was detected in samples other than sputum during the entire nucleic acid detection process. It can be seen that the virus in this patient mainly existed in the lower respiratory tract, and the viral load in the nasopharynx was low. It may also be one of the reasons accounting for its several false-positive RT-PCR tests and not infection of her close contacts. Similarly, no virus detected in anal swabs and stool samples proved that the virus did not infect the digestive tract, so the patient did not experience symptoms such as abdominal pain and diarrhea. But whether the distribution of the virus affects the spread of the virus needs more studies.

Most notably, although the patient was undiagnosed by eight nucleic acid tests in this case, the patient’s clinical manifestations, imaging examinations, and other laboratory tests showed that he had SARS-CoV-2 infection. For such multiple “false positive” cases, specific antibody detection of SARS-CoV-2 in the patient’s blood can be used as a supplement to nucleic acid detection. In the latest “Diagnosis and Treatment of COVID-19 (Trial Version 7)”, it is mentioned that the serological antibody testing is used as the basis for diagnosis. COVID-19 is a new infectious disease and is throughout the course of disease same as SARS. The specific IgM of SARS-CoV mainly exists in the early and middle stages of infection and specific IgG mainly exists in the middle stage of infection and recovery period [12, 13]. Because SARS-CoV-2 and SARS-CoV belong to the same family, the detection of IgM and IgG antibodies against SARS-CoV-2 may cause the presence of SARS-CoV-2 in patients with negative nucleic acid tests. Therefore, the negative result of several nucleic acid tests may be due to the high specific IgM in the early stages of onset. The high specific IgM makes the SARS-CoV-2 low load in the body and amount of virus insufficient in the sample, leading to RT-PCR result is negative. But the virus does exist in the patient and is infectious at early stage. Considering that the production of specific antibodies takes time, and antibody detection is easily affected by rheumatoid factor, non-specific IgM, autoantibodies in blood samples, the RT-PCR result could be false positive. So even if the antibody is positive, continue to take samples to detect the viral nucleic acid. With the outbreak in many places around the world, more and more infected people, like the patients in this study, may have no obvious clinical symptoms or even no clinical symptoms after infection[14], and have negative or single-target positive RT-PCR results[4]. For such infected persons, it is necessary to combine the efficient SARS-CoV-2 capture sequencing to detect viral nucleic acids, helping clinical diagnoses. In addition, because the RNA virus is susceptible to mutation, sequencing of pathogenic nucleic acid genomes from samples of asymptomatic and occult infected patients is also conducive to studying the virus mutations in the pathogenic genes providing a basis for subsequent virus tracing and epidemiological investigations.

As the virus spreads, research data from Chen et al [15] showed that the initial symptoms of newly infected patients seemed to be more subtle, and the virus may lie in asymptomatic carriers for a long time. Similarly, our study also showed that it is also difficult to detect infections due to the reason that the clinical symptoms of the COVID-19 in the early stage of infection were mild or even absent. Such unknown infection has caused 79% of the overall infection [16], and is the main reason for the widespread spread of this virus. The patient in this case was isolated in a timely manner after her colleague was diagnosed, reducing the risk of transmission. Therefore, it can be seen that the follow-up of COVID-19 patients’ contacts or experience areas is necessary to control the disease. Considering asymptomatic and mildly symptomatic infections, it is recommended that contacts of confirmed patients should be screened and tested for viral nucleic acid regardless of the presence or absence of symptoms, and preventive use of effective antiviral drugs for asymptomatic patients to reduce viral load and reduce the risk of virus transmission from asymptomatic carriers. Such measures may help control the spread of COVID-19. Similarly, it is not yet possible to determine whether patients have virus or are still infections in the recovery period although RT-PCR tests are negative. Therefore, it is recommended to increase the number of tests and the interval between each test.

## Conclusion

We report the epidemiological history and clinical information of a patient with negative (or single-target positive) SARS-CoV-2 infection with multiple RT-PCR tests. The present case indicates that mild patients with virus-free or low-load in the upper respiratory tract may mainly cause several RT-PCR results false negatives during the diagnosis. Therefore, it is recommended to strengthen the detection and isolation of asymptomatic or mild patients to avoid the wide spread of COVID-19.

## Data Availability

All data included in this study are available upon request by contact with the corresponding author.

## Ethics

This study was approved by the ethics commissions of the participating hospitals, with a waiver of informed consent.

## Figure legends

**Figure 1.** The patient’s Chest CT images. A: CT images on Feb 12; B: CT images on Feb 15.

**Figure 2.** Body temperature of the patient.

**Figure 3.** Chest Radiographs. Image on Feb 28

